# Estimating SARS-CoV-2 reproduction number by infection location in Japan

**DOI:** 10.1101/2021.04.13.21255296

**Authors:** Junko Kurita, Takahide Hata, Tamie Sugawara, Yasushi Ohkusa, Atsuko Hata

## Abstract

**Background:** COVID-19 infectiousness might differ by infection location. Nevertheless, no such study of infectiousness has been reported.

**Object:** The study objective was estimation of the reproduction number by infection location.

**Method:** Patients who infected no one were ignored because their reliability might be lower than that of patients who infected more than one person. On the assumption that the histogram follows an exponential distribution, we estimated the reproduction number from the histogram of the number of people infected by the same patient.

**Results:** Night entertainment venues showed the greatest infectiousness, followed by facilities for elderly people and hospitals. Nursery schools and workplaces were followed by homes, with the lowest infectiousness.

**Discussion and Conclusion:** Countermeasures under the second declaration of emergency status targeted restaurants. However, infectiousness at restaurants was not high. Comparable to those of universities and karaoke, and not significantly different from homes: the least infectious location studied.

## Introduction

Since the emergence of COVID-19 in December, 2019 in Wuhan, China, reproduction numbers have been estimated several times. Some of the earliest studies conducted in Wuhan [1–3] estimated *R*_0_ for COVID-19 as 2.24–3.58. Even in Japan, early research [4] estimated *R*_0_ as 2.049 (95% confidence interval (CI) [2.403, 2.557]). However, these reproduction numbers were for whole populations. Reproduction numbers by location of infection are less known, but infectiousness probably differs among infected places. For instance, countermeasures under the second emergency status declaration on January 7, 2021 clearly stipulate that restaurants close earlier than eight o’clock p.m. This policy was based on an inference that infectiousness at restaurants was higher than at other areas. By contrast, nursery schools and schools were not required to close as a countermeasure, although they had been closed under the first emergency declaration from April 8 to May 24, 2020. The objective of this study was confirmation of differences in infectiousness by infection location.

A study conducted to estimate infectiousness in the earlier stage of the outbreak in Japan included patients who were not reported as having infected someone [5]. They estimated a very small reproduction number, 0.6, as of the end of February in Japan. Although they did not designate it as *R*_0_, they referred to it as the average number of secondary infections. Such a low number indicates that the outbreak of COVID-19 was self-limited. Therefore, any intensive infection control such as school closure or restriction against going out is expected to be unnecessary. The authors of that report apparently misunderstood the meaning of patients who were not reported as having infected someone. They might have been severely underestimated at that time. Therefore, people they infected might have been found and reported. Alternatively, investigation of them cannot simply reveal who had been infected by them. Therefore, we proposed another method of estimating infectiousness that excluded information of patients who were reported as not having infected anyone [6].

When we applied our proposed procedure for the present study to data obtained from an earlier study, we obtained a figure of 4.4273. Its 95% CI was [3.6000, 5.3364]: more than six times greater than the original estimate. That finding was comparable to our results obtained for infections from adults to elderly people and from elderly people to adults. They apparently underestimated *R*_0._ Therefore, the chosen infection-control policy was misguided, with insistence on contact tracing.

## Method

We adopted a similar method to estimate infectiousness by location of infection, as in our earlier study, which investigated infectiousness by age of the infected person and age class inferred from the infection source [6].

We chose to examine major places where people were being infected: homes, hospitals, facilities for elderly people, workplaces, schools, nursery schools, universities, restaurants, night entertainment venues, and karaoke. For this study, the School category does not include nursery schools or universities, but does include kindergartens, elementary schools, junior high schools, and high schools. In addition, the Restaurant category does not include night entertainment venues or karaoke.

Let *x*_*i,j*_ represent the number of cases in which *j* patients were infected secondarily in place *i*. Because we do not know the probability by which a patient infected one person, the probability that a person infected two or more people was assumed to follow an exponential distribution as *p*_*i*_, *p*_*ij*_^2^, *p*_*ij*_^3^, and so on. Then *R*_*ij=*_ *p*_*ij*_*+*2*p*_*i*_^2^*+*3*p*_*ij*_^3^*+…*=Σ_*k*=1_ *k p*_*ij*_^*k*^*=p*_*ij*_/(1-*p*_*ij*_)^2^.

We observed an estimator of *p*_*i*,_ as *x*_*i*,1_/*N*_*i*_, where *N*_*i*_, represents an unknown total number of age class *i* patients who were infected by age class *j* patients. Similarly, *p*_*i*_^2^ was estimated in general as *x*_*i*,2_/*N*_*i*_ and *p*_*i*_^*m*^ = *x*_*i,m*_/*N*_*i*_. By log transformation, we have *m log p*_*i*_*= log x*_*i,m*_ *– log N*_*i*_ (*m*=1, 2,*…M*) where *M* stands for the maximum number of secondary infections. Therefore, we obtain an estimator of *p* as an estimated coefficient of regression of *log x*_*i,m*_ on *m* using ordinary least squares method. In addition, *N*_*i*_^*\**^ was obtained from an exponential transform for the estimated constant term.

The confidence interval (CI) of *R*_*i*,_^*^ was obtained using a bootstrapping procedure for the distribution of {*x*_*i,m*_ (*m*=1, 2, *…L* _*i*_)}, where *L* _*i*,_ stands for the maximum number of non-zero secondary infection [4]. In addition to usual bootstrapping, we conducted it with special consideration for the case of *x*_*i,m*_*=*0 (*m=*1, 2, *…L* _*i*_). These cases were ignored in estimation despite including much information. We bootstrapped for the distribution of {*x*_*i,m*_*+*1 (*m*=1, 2, *…L*_*i*_)} and produced an estimate using max[0.001,{*x*_*i,m*_*+*1}^*b*^-1](*m*=1,2, *…L*_*i*_), where superscript *b* denotes a bootstrapped series and 0.001 was a small number instead of 0.

Based on the *j*-th bootstrapped distribution {*x*_*i,m*_ (*m=*1, 2,…)}^*j*^, we can obtain *R*_*i,j*_^*^. We repeated this procedure one million times, thereby obtaining one million bootstrapped *R*_*i,j*_^*^. We sorted these variables. The duration from *R*_*i*,25000_^*^ to *R*_*i*,975000_^*^ is expected to be 95% CI of *R*_*i*_^*^.

All information used for this study was obtained from reports of the Ministry of Health, Labour and Welfare [7] and local governments. The study period extended from January 15, when the initial case was detected in Japan, to the end of July.

### Ethical considerations

All information used for this study has been published elsewhere [7]. There is therefore no ethical issue related to this study. We inferred significance at the 5% level.

## Results

Through the end of July, 36,431 patients had been confirmed in Japan. From those, after excluding asymptomatic cases, cases of people presumed to have been infected in foreign countries, and cases for which no onset date was available, we were left with 30,780 cases. Of those cases, after excluding cases for which the infection source was unknown, and cases for which the age of patients and sources of infection were unavailable, we were left with 5383 cases. Of those, 4886 cases were identified as infection sources. These 4886 cases were analyzed for this study.

Figure 1 presents a histogram of cases by the number of secondary infections at home. Figure 2 depicts infection cases related to hospitals, facilities for elderly persons, or workplaces. It is noteworthy that three cases showing 20 secondary infections in a hospital represented more than 20 secondary infections. Figure 3 portrays those in schools, nursery schools, and universities. Figure 4 depicts those in restaurants, night entertainment venues, and karaoke.

**Figure 1:**
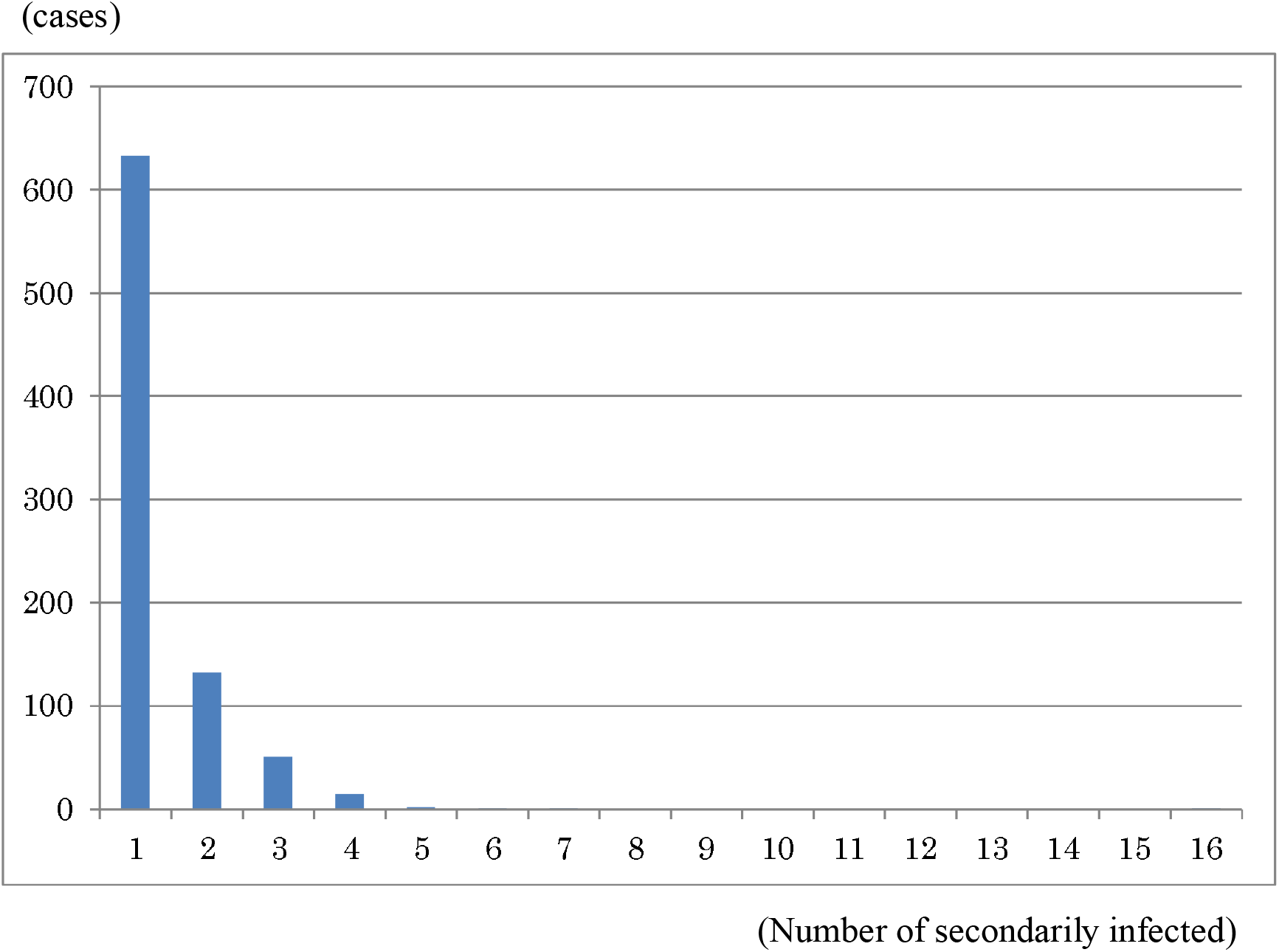
Histogram showing the numbers of infected cases at home. Note: Bars represent numbers of people infected at home.

**Figure 2:**
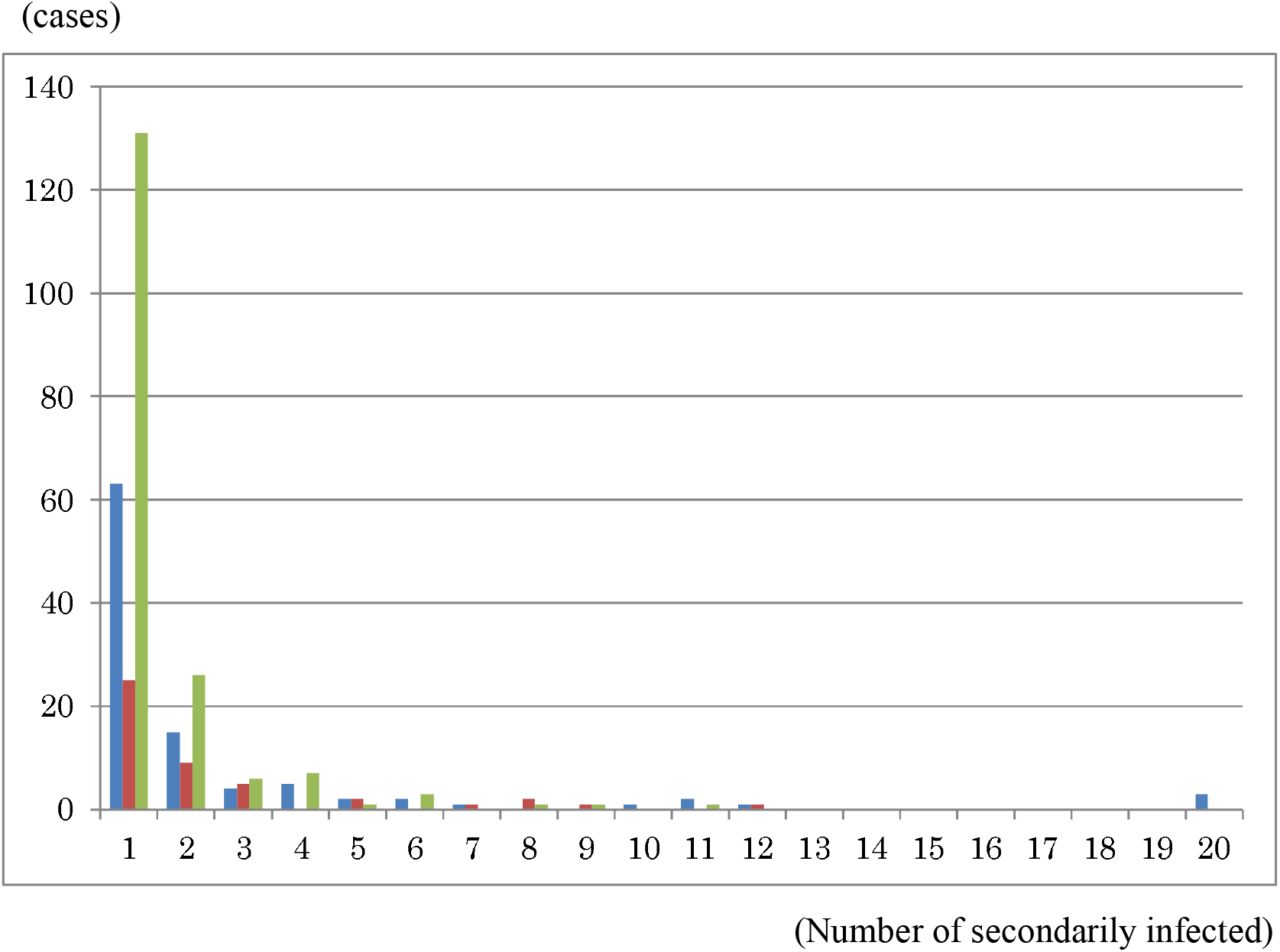
Histogram showing numbers of people infected at hospitals, facilities for elderly people, and workplaces. Note: Blue bars represent the number of the infected cases at hospitals. Orange bars represent those at facilities for elderly people. Gray bars represent those at workplaces. Infections at hospitals include cases in which 21, 34, or 57 were secondarily infected. In the figure, these three cases were added together as 20 secondarily infected.

**Figure 3:**
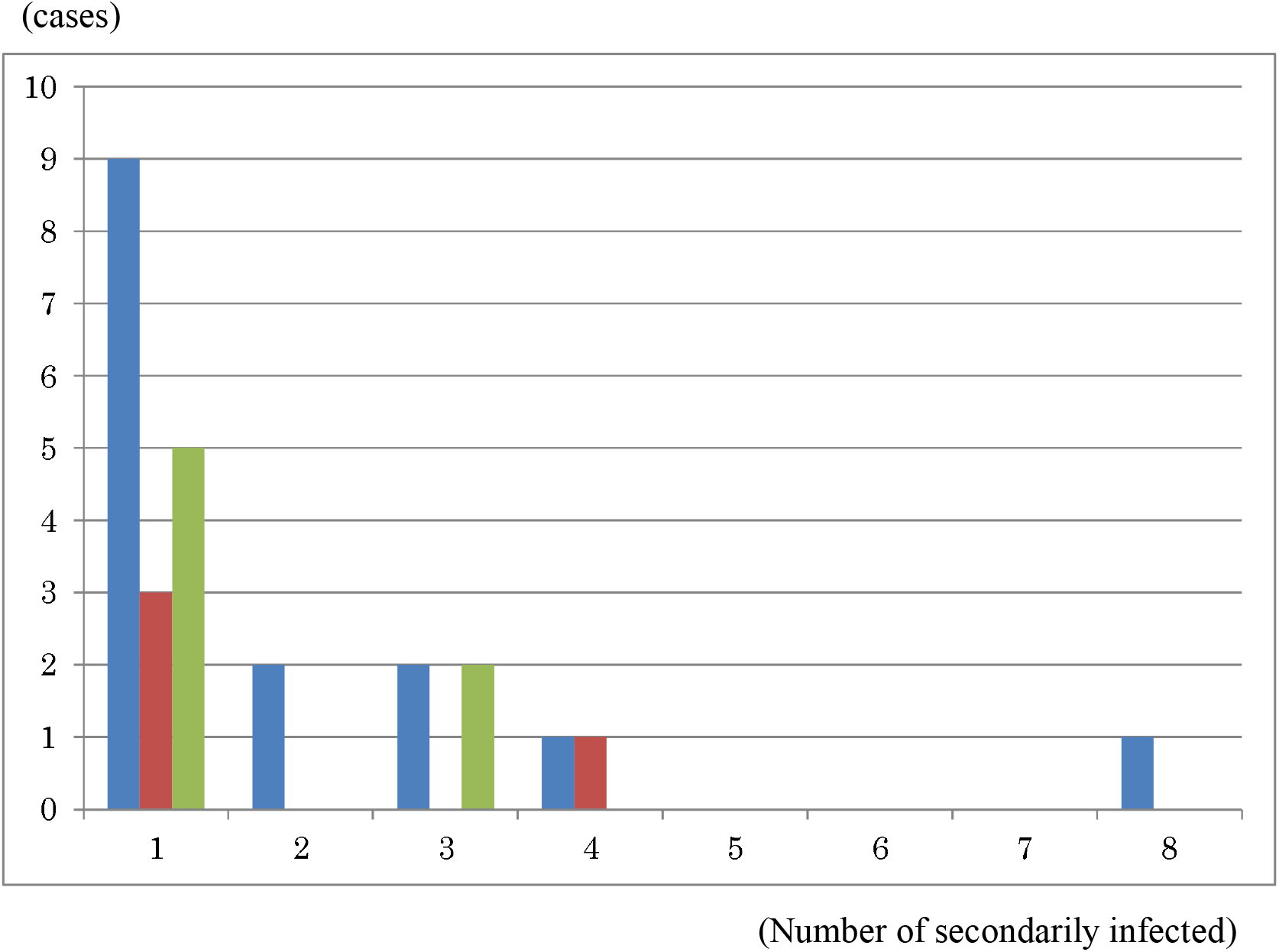
Histogram showing the numbers of people infected at schools, nursery schools, and universities. Note: Blue bars represent the number of the infected cases in school, Orange bars represent those at nursery school. Gray bars represent those at university. Schools do not include nursery schools or universities, but include kindergartens, elementary, junior high, and high schools.

**Figure 4:**
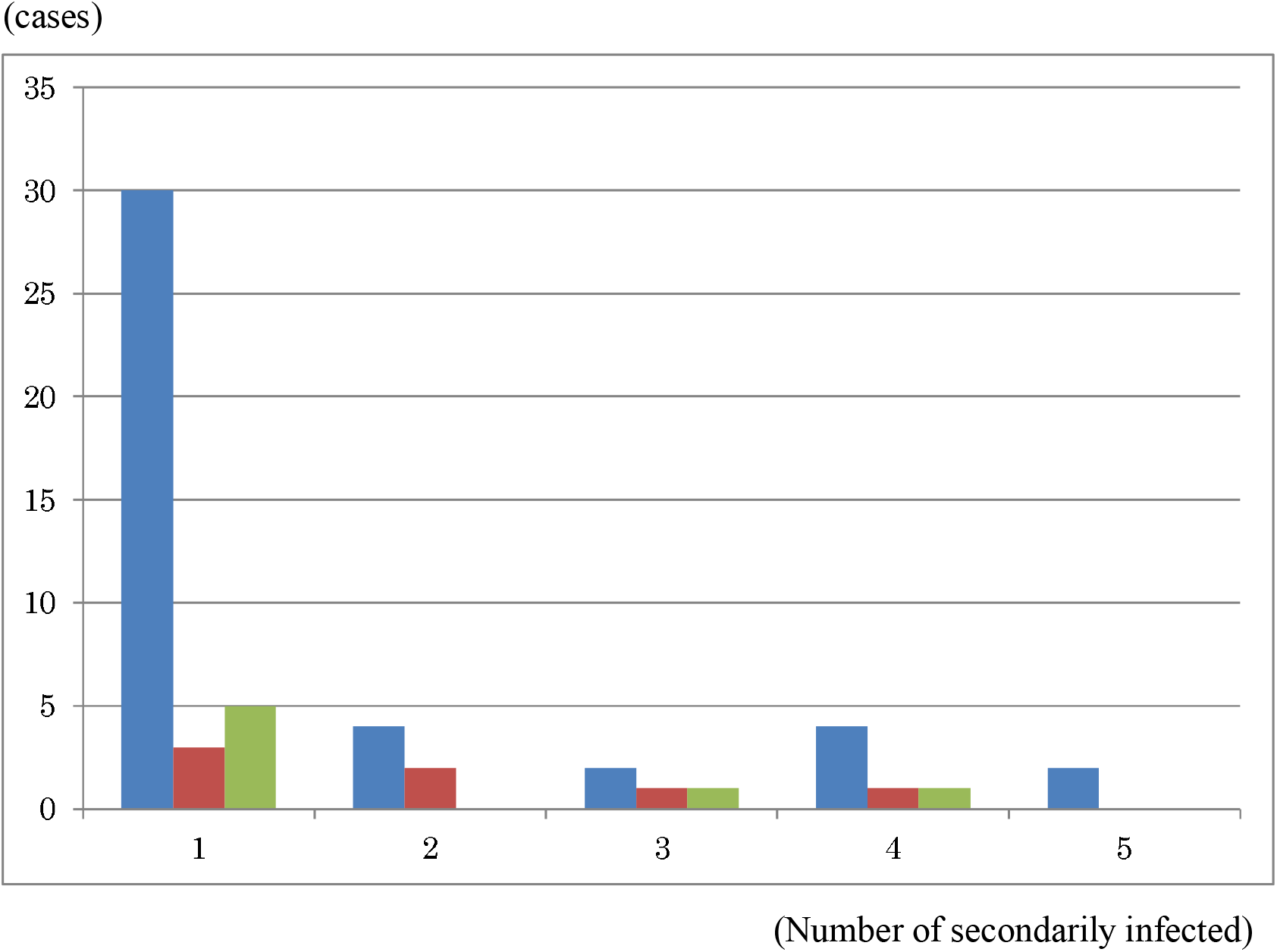
Histogram showing numbers of people infected at restaurants, night entertainment venues, and karaoke. Note: Blue bars represent the number of people infected at restaurants. Orange bars represent those infected at night entertainment venues. Gray bars represent those infected at karaoke. Restaurants do not include night entertainment venues or karaoke.

Estimation results of *R*_*i*_ are presented in Table 1. Regarding median values, hospitals were found to be the highest, followed by universities and facilities for elderly people. Night entertainment venues were the lowest, followed by nursery schools, schools, and workplaces.

**Table 1:**
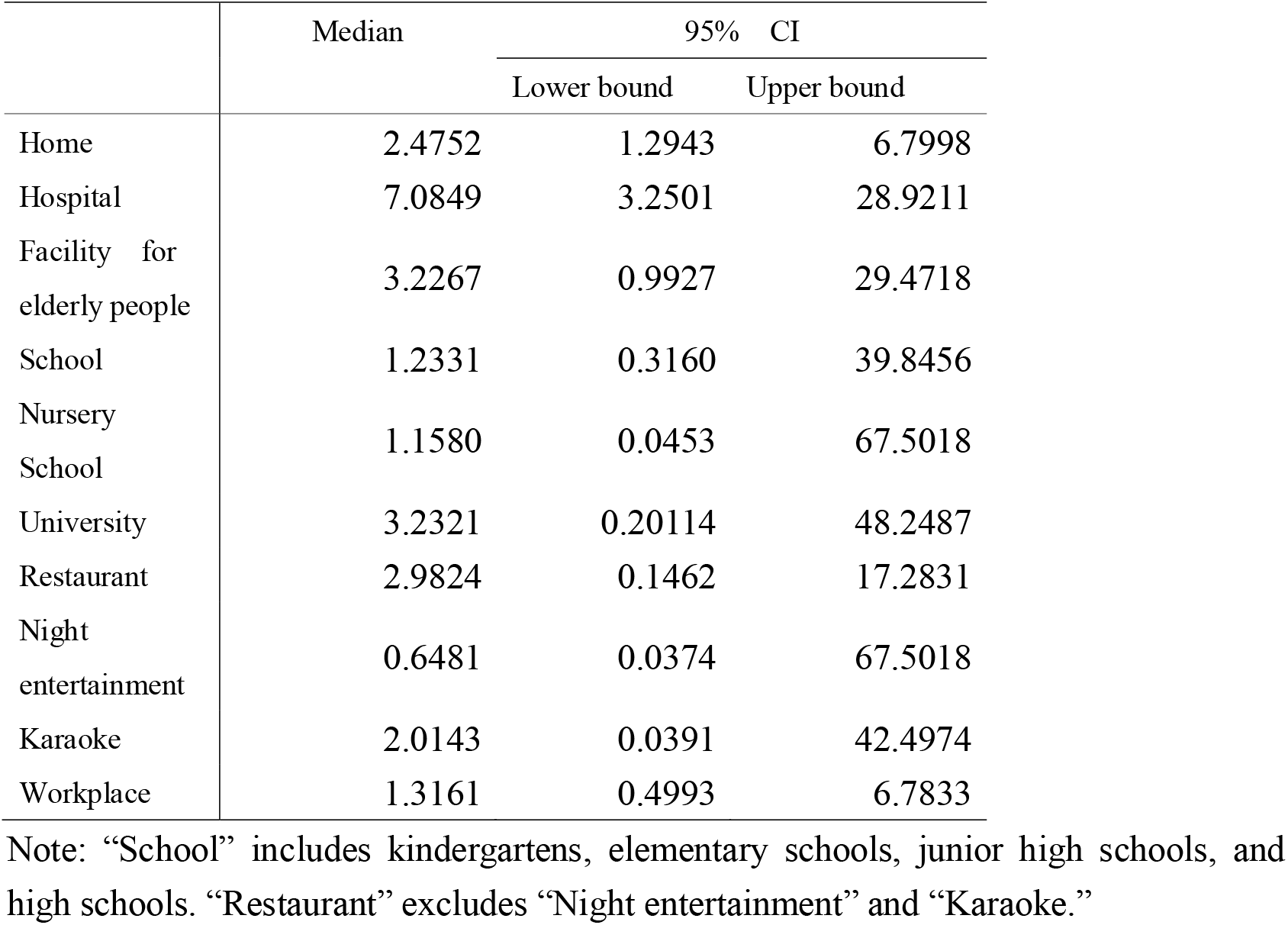
Estimated results of effective reproduction number by infection location

However, except for homes and hospitals, the lower bound of 95% CI of all other sites was less than one. In other words, their infectiousness was not significantly different from one. Therefore, their infectiousness at hospitals and homes was considerably higher than one.

## Discussion

We used a procedure to estimate the case distributions among numbers of infected cases developed in our earlier study [6]. Although infected cases or unlinked cases for which the infection source was unknown represented a majority of cases, the procedure we used ignores information those cases because it was less credible. However, information about patients who were reported as having infected someone was more reliable than others because, at least, they had been investigated by public health authorities.

Results demonstrated that the estimated infectiousness at hospitals and homes was significantly greater than one. Infectiousness at facilities for elderly people was marginally higher than one. Infectiousness at the other considered places was not significantly higher than one. Particularly, the estimated infectiousness in restaurants was not high. Therefore, rather than restaurants, countermeasures for COVID-19 should specifically examine hospitals, some other considered places, or homes.

It is noteworthy that infectious areas found from the present study do not represent a hot spot at which numerous people were infected. The total number of people infected in a type of place represents the product of infectiousness and people who are infectious visiting and staying at a place. For example, although infectiousness at homes was less than at other places, a huge number of patients stayed at home and shared contact with family members. For those reasons, one would expect that the number of people infected at home would be quite larger than at other places: and it was. When interpreting the obtained results, one must be reminded that infectiousness represents an average number of secondarily infected people per infectious person.

We have examined advanced bootstrapping procedures with special consideration for some particle numbers of secondary infection recording zero cases. For estimation in the present study, information about the number of secondary infections was ignored because log transformation of the number of cases was used. However, the likelihood of one case at a particular number of secondary infections actually leading to zero cases was probably less but an almost comparable likelihood to that of one case at a particular number of secondary infections actually recording one case in a bootstrapping procedure. Therefore, we treat those numbers of secondary infections recording zero cases with special consideration.

The present study has some limitations. First, because infectiousness in all places were not significantly different as results, data might be insufficient to do our procedure. When we accumulate the data, it might be solved partially.

Second, because of data limitations, we cannot analyze characteristics such as those of patients or hospital staff, residents or staff at a facility for elderly persons, or students and teachers at a school. For example, infectiousness among students in school or among kids in nursery school, or of medical staff to patients are probably very important factors to control the outbreak. To resolve that difficulty to some degree, data accumulation is expected to be necessary in the near future.

Thirdly, seasonality of infectiousness might be fundamentally important, as it has come to be for influenza. Because data used for this study were accumulated through July, we are unable to evaluate them. In winter, data must also be analyzed similarly. Risk related to location must be evaluated.

## Conclusion

This study demonstrated that effective reproduction numbers at restaurants were not high. Results show that they were comparable to data from universities or karaoke and were not significantly different from data related to infection at home: the least infectious place. Therefore, countermeasures taken under the second emergency status declaration targeting infection at restaurants might not be based on evidence. We can find no significant difference in infectiousness among the places considered.

The present study is based on the authors’ opinions: it does not reflect any stance or policy of their professionally affiliated bodies.

## Data Availability

Japan Ministry of Health, Labour and Welfare. Press Releases.

https://www.mhlw.go.jp/stf/newpage_10723.html

## Acknowledgments

We acknowledge the great efforts of all staff at public health centers, medical institutions, and other facilities who are fighting the spread and destruction associated with COVID-19.

## Ethical considerations

All information used for this study was collected under the Law of Infection Control, Japan and published data was used. There is therefore no ethical issue related to this study.

## Competing Interest

No author has any conflict of interest, financial or otherwise, to declare in relation to this study.

## Notes

### Competing Interest Statement

The authors have declared no competing interest.

### Funding Statement

The author(s) received no specific funding for this work.

### Author Declarations

All information used for this study has been published. There is therefore no ethical issue related to this study.

### Summary of Updates

I regret my mistakes about authors' affilication. I fixed thiese mistakes in thie version.

